# Deep learning clarifies association of osteoporosis risk with bone metastasis in premenopausal women after surgery for early-stage breast cancer: a multicenter retrospective cohort study

**DOI:** 10.1101/2025.03.23.25324493

**Authors:** Hirokazu Shimizu, Nobumoto Tomioka, Akira Iwata, Chinatsu Koganezawa, Tomohiro Oshino, Kenichi Watanabe, Mitsugu Yamamoto, Hiroaki Hiraga, Masatake Matsuoka, Yoichi M Ito, Shinya Tanaka, Masato Takahashi, Norimasa Iwasaki

## Abstract

**Background:** Adjuvant use of bone-modifying agents (BMAs) to early-stage breast cancer (eBC) aims to maintain bone density, leading to prevention of bone metastasis (BM) in postmenopausal women; its mechanism remains unknown. Clinically, one-quarter of premenopausal women develop osteopenia. To this, deep learning (DL) enables to evaluate the risk of osteoporosis (RO) using perioperative chest radiography. The aim of the study was to clarify the association of RO with BM after eBC surgery and perform stratified analyses by menopausal status.

**Methods:** In this multicentre retrospective cohort study, we enrolled 785 women who underwent surgery for estrogen receptor-positive eBC between 2007 and 2019, excluding those with adjuvant BMAs. Perioperative chest radiography was interpretated into the DL-producing RO score (0-1), classifying them into 359 women with low risk of osteoporosis (LRO) and 426 women with high risk (HRO). The 5-year BM-free survival (BMFS) was compared.

**Findings:** Univariate stratification by menopausal status for the enrolled women (median [interquartile range] age, 56 [46-66] years) revealed: the number of premenopausal women, 325; HRO (23.1% [75/325]) in premenopausal women was associated with low BMFS (hazard ratio [HR] 2.37, 95% confidence interval [CI]; 1.23-4.57, p<0.01), whereas no association was observed in the others (p=0.78). Multivariate analyses identified RO as a poor prognostic factor in the enrolled women (HR 3.08, 95%CI; 1.23-7.47, p=0.01). Conduction rate of bone density tests was lower in premenopausal women (31/325 vs. 317/460, p<0.01).

**Interpretation:** RO is an independent prognostic factor for BM in women with eBC. Our study revealed association between RO and BM, specifically in premenopausal women, whose bone health is poorly studied. Expanded indications of adjuvant BMA are suggested for at-risk premenopausal women.

**Funding:** None.

## Introduction

Breast cancer remains the second leading cause of cancer death among women^1^. As approximately 645000 premenopausal breast cancer cases were diagnosed worldwide in 2018^2^,the incidence of breast cancer has been increased, rising by 1 % annually during 2012-2021^1^. In addition, breast cancer in younger women is generally aggressive and their prognoses are relatively poor^3^; patients with breast cancer aged 40–60 years are prone to bone metastasis (BM)^4,5^. As the most common first site of distant spread is the bone in breast cancer,^6^ a biomolecular research study revealed that bone environments invigorate metastatic seeds for further dissemination.^7^ Thus, preventing bone metastasis is a crucial therapeutic strategy.

Randomised controlled trials (RCTs) suggest that adjuvant bone-modifying agents (BMAs) can improve bone metastasis-free survival (BMFS) in postmenopausal women with early-stage breast cancer (eBC).^8-10^ The precise mechanism by which BMAs regulate the occurrence of BM is not fully understood, and it remains unknown whether it is applicable to younger population. An original concept of the adjuvant BMA use is the maintenance of bone density, thereby preventing osteoporosis.^9,11,12^ In clinical settings, approximately one-quarter or more of premenopausal or perimenopausal women might suffer from osteopenia.^13-15^ There is a certain proportion of premenopausal eBC women with osteopenia, but a bone density test in perioperative periods is often omitted. Thus, direct evaluation of osteoporosis or osteopenia using DXA in premenopausal women with eBC is challenging.

Rapid advances in deep learning (DL) have made it possible to accurately assess the risk of osteoporosis (RO) in patients only by using chest radiography.^16-18^ This suggests that the assessment can be performed in patients who have had perioperative chest radiography evaluation for surgery under general anaesthesia.^19^ Based on the concept of adjuvant BMA use, we hypothesised that osteopenia contributes to early recurrence of bone metastases in premenopausal women. The aim of this study by utilizing DL was 1) to reveal the association between RO and BM in women with eBC, and 2) to perform a stratified analysis by menopausal status.

## Methods

### Study design and participants

In this multicentre retrospective cohort study, we recruited patients based on the surgical records of breast cancer between 2007 and 2019 (Figure1). We recruited 862 patients who underwent surgery for clinical stages 2–3, were estrogen-positive, and had human epidermal growth factor receptor2 (HER2)-negative breast cancer. Seventy-seven patients were excluded because they were males, had bilateral breast cancer, did not have perioperative chest X-ray, or had adjuvant use of BMA owing to osteoporosis (T-score of -2.5 or low). A total of 785 women were enrolled in this study.

**Figure 1.**
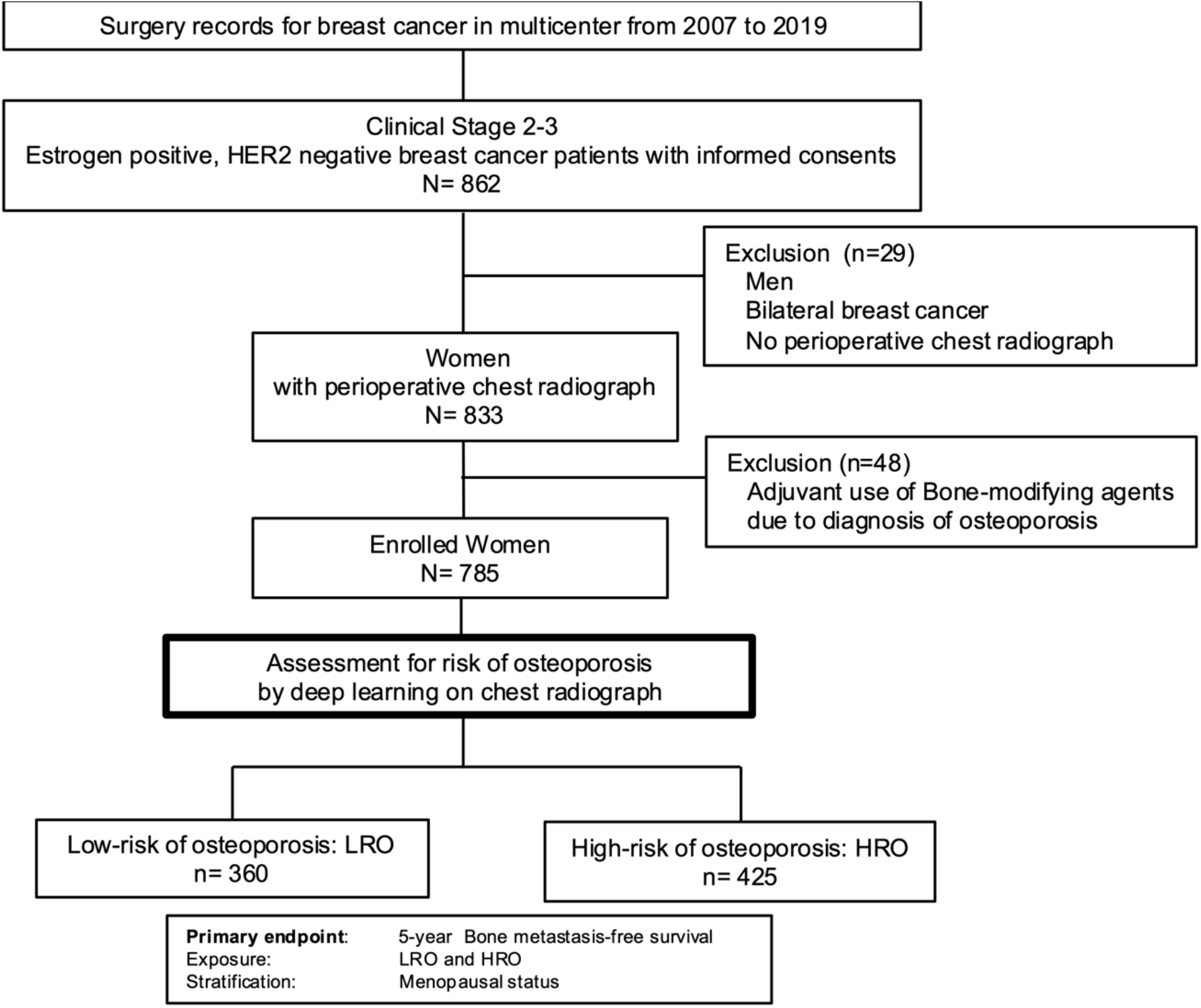
Study design

This study was approved by the local ethics committees of Hokkaido University Hospital (022-0118) and the participating hospital, NHO Hokkaido Cancer Center. Informed consent was obtained from all participants who agreed to the use of their data. Clinical trials were not included in this study. The study was conducted in accordance with the principles of human ethics outlined in the Declaration of Helsinki.

### Procedures

The enrolled women were classified by RO estimated using the deep learning application, OPSCAN,^16,17^ which yields risk score as a continuous variable [0-1]. As the output of the model was to predict the T-score, a higher risk score correlated with a lower T-score.^17^ The RO score was used for classification: 359 women with low-risk osteoporosis (LOR) and 426 women with high-risk osteoporosis (HOR). In our original cohort, electronic medical records were used to ascertain the clinicopathological characteristics, including perioperative chest radiography and bone density tests. The following primary treatment data were collected: dates of surgery, radiotherapy, perioperative chemotherapy, and endocrine therapy.

### Outcomes

As the primary outcome, the 5-year bone metastasis-free survival (5-year BMFS) was compared. A Cox proportional hazards model was used to identify the independent prognostic factors. A stratified analysis was performed according to menopausal status which affects the bone environment. Definition of menopause was based on the US guideline^20^.

### Statistical analysis

Categorical variables were evaluated using the chi-square test for two or more groups, whereas continuous variables were analysed using an independent t-test. The normality of the distribution of patient age was assessed using the Shapiro-Wilk test. Survival rates were visualised using the Kaplan–Meier method, and differences were investigated using the log-rank test. A Cox proportional hazards model was used to identify the independent prognostic factors. A stratified analysis was performed according to menopausal status which affects the bone environment. The Pearson correlation coefficient was used to measure the linear correlation between the two sets of data. All statistical analyses were performed using JMP Pro version 14 (SAS Institute Inc., Cary, NC, USA). All tests were two-sided and *p<*.*05* was considered statistically significant.

## Results

### Characteristics of the study cohort

The baseline characteristics of the enrolled women (median [interquartile range (IQR)] age 56[46-66]) are shown in Table1. Women in HRO were older during surgery than LOR (median [IQR] age, 47 (43-53) vs 64 (59-70), p<0.01). The rate of stage 3 disease did not differ between the HRO and LRO groups (101/425 vs. 96/360, p=0.33). The rate of perioperative chemotherapy was lower in the HRO than in LRO groups (198/425 vs. 246/360, p<0.01). The rate of endocrine therapy did not differ between the two groups (398/425 vs. 337/360, p=0.96). In the premenopausal women, rate of tamoxifen monotherapy was higher in HRO than LRO groups (57/75 vs 153/250, p=0.02)

**Table 1.** Characteristics of the cohorts.

| Characteristics | Low risk of osteoporosis<br>n=360 | High risk of osteoporosis<br>n=425 | p value |
| --- | --- | --- | --- |
| Age, median(IQR) | 47 (43-53) | 64 (59-70) | <0.01 |
| Body mass index, kg/m<sup>2</sup> (SD) | 24.1(4.9) | 23.9(4.6) | 0.44 |
| Menopause (%) | 110(30.6) | 350(82.4) | <0.01 |
| Stage |  |  | 0.33 |
| 2 | 264 | 324 |  |
| 3 | 96 | 101 |  |
| Nodal involvement (%) | 228(63.3) | 250(58.8) | 0.22 |
| Radiation therapy (%) | 171(47.5) | 191(44.9) | 0.43 |
| Perioperative chemotherapy (%) | 246(68.3) | 198(46.6) | <0.01 |
| Endocrine therapy (%) | 337(93.6) | 398(93.6) | 0.96 |
| Risk of osteoporosis score [0–1] | 0.04±0.03 | 0.44±0.24 | <0.01 |

IQR; interquartile range, SD: standard deviation, osteoporosis risk is the highest for score of 1

In the enrolled women, bone was most common as the first distant recurrence site (HRO, 48 of 425 women[14.8%], LHO 29 of 360 women [8.1%]), followed by liver (HRO, 11 of 425 women[3.4%], LHO 18, of 360 women [5.0%]), lung (HRO, 13 of 425 women[4.0%], LHO, 7of 360 women [1.9%]), and brain (HRO, 3 of 425 women[0.9%], LHO 3 of 360 women [0.8%]). 5-year follow-up rate was 97.8% (768/785).

### The 5-year BMFS ratio stratified by menopausal status

Univariate analysis showed that HRO was not associated with BM among the enrolled women (log-rank test, p=0.05) (Figure 2a). As stratification was conducted by menopausal status, premenopausal women (n=325) were HRO (23.1%, 75/325) with a lower 5-year BMFS (p<0.01) (Figure 2b). There was no significant difference between the two groups in post-menopausal women (n=460) (p=0.78) (Figure 2c). Menopausal status stratified the recurrence of BM. In addition to menopausal status, age, body mass index, and perioperative chemotherapy, which are known risk factors for cancer and osteoporosis^21-24^, we included the above variables in our statistical models.

**Figure 2.**
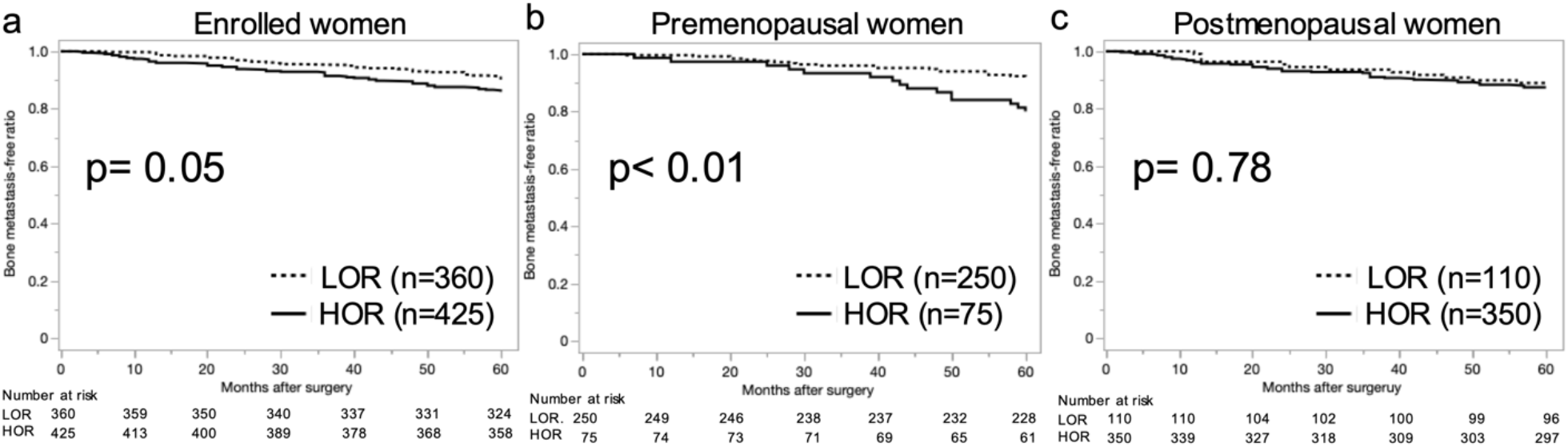
Univariate analyses stratified according to the menopausal status Survival analysis representing the proportion of bone metastasis-free patient-based risk of osteoporosis score*s* in a) all enrolled women, b) premenopausal women, and c) postmenopausal women. LRO; low risk of osteoporosis, HRO: high risk of osteoporosis

### Multivariate analysis

Cox proportional hazard models adjusted by age, body mass index, menopausal status, and the presence of perioperative chemotherapy demonstrated that RO score (hazard ratio [HR] 3.08, 95% confidence interval [CI]; 1.23–7.47, p=0.01) and clinical stage (HR 3.05, 95%CI; 1.98– 4.73, p<0.01) were independent prognostic factors (Table 2). The models also demonstrated that RO score was an independent poor prognostic factor in premenopausal women (HR 7.53, 95%CI: 1.54–28.61, p<0.01), but not in postmenopausal women (HR 1.97, 95%CI; 0.70–5.57, p=0.20)

**Table 2.** Cox proportional models for recurrence of bone metastasis in women after definitive surgery for early-stage breast cancer.

|  | Hazard ratio | 95% confidence interval | *P value* |
| --- | --- | --- | --- |
| Risk of osteoporosis score |  |  |  |
| Enrolled women | 3.08 | 1.23–7.47 | 0.01 |
| Premenopausal women | 7.53 | 1.54–28.61 | <0.01 |
| Postmenopausal women | 1.97 | 0.69–5.57 | 0.20 |
| Stage 3 |  |  |  |
| Enrolled women | 3.05 | 1.98–4.73 | <0.01 |
| Premenopausal women | 3.14 | 1.58–6.23 | <0.01 |
| Postmenopausal women | 2.82 | 1.58–5.00 | <0.01 |

adjusted by age, body mass index, menopause, perioperative chemotherapy

### bone density test and risk of osteoporosis

The rate of perioperative bone density tests in the enrolled women was 44.3% (348/785). Importantly, the rate in premenopausal was 9.5% (31/325), whereas that in postmenopausal women was 68.9% (317/460) (p<0.001) (Figure3a). In postmenopausal women, the risk of osteoporosis and T-score were correlated (n=317, r=-0.61, p<0.01) (Figure3b). A formula for predicting bone metastasis within 5 months was generated using independent prognostic factor staging and the risk of osteoporosis (Figure 3c) to ensure the validity of the risk of osteoporosis in premenopausal women. There was moderate performance in predicting bone metastasis within 5 years of surgery in premenopausal women with an area under the receiver operating characteristics of 0.71 (Figure 3d).

**Figure 3.**
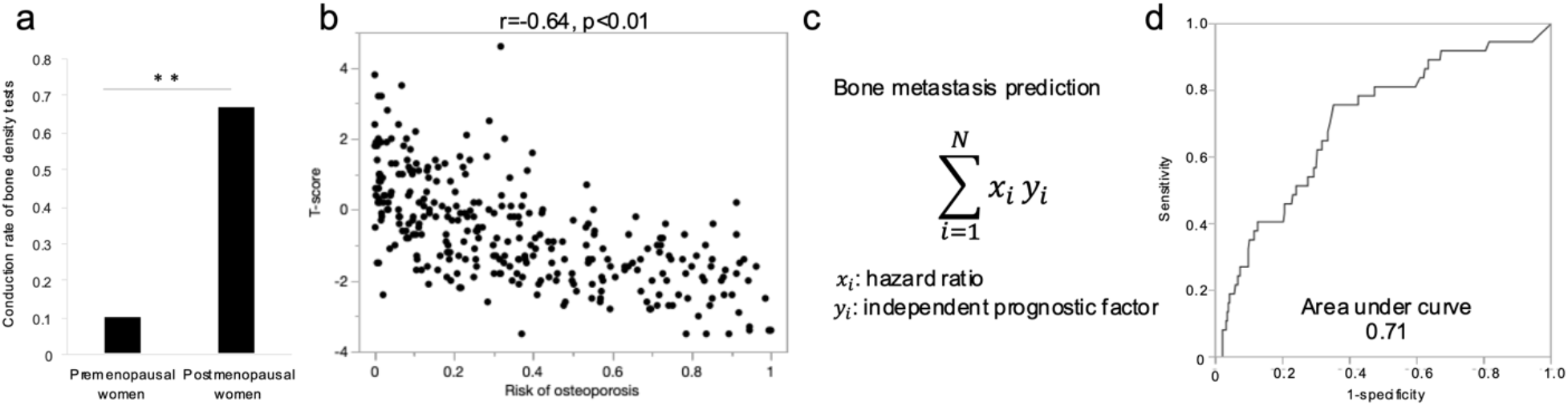
Validity of the risk of osteoporosis a) Conduction rate of bone density tests in premenopausal and postmenopausal women. b) Correlation between T-scores and risk of osteoporosis in postmenopausal women. c) Formula for predicting bone metastasis within five years after surgery for early-stage breast cancer. d) Receiver-operating characteristic curves of the formula for predicting bone metastasis in premenopausal women.

## Discussion

We found RO score as an independent prognostic factor for bone metastasis in women with eBC, revealing a clear association between RO and BM, especially in premenopausal women. Current clinical settings have allowed physicians to consider the adjuvant use of BMA only in postmenopausal women with eBC. Our data showed that the conduction rate of the bone density test was lower than 10% in premenopausal women. Further studies should prospectively confirm the association of T-scores in premenopausal women with eBC with BM recurrence, along with the clinical benefit of adjuvant BMA use.

Recent biomolecular research has focused on whether conditions that alter depending on the age of the host environment can affect the metastatic recurrence of secondary lesions.^25,26^ In the lung, one of the most frequent secondary sites for lesions, age-induced reprogramming of lung fibroblast promotes tumour cell activation and proliferation.^25,26^ The bone is another well-known and common secondary site where the activities of osteoclasts are age-dependent.^27-29^ Osteolytic metastases, such as in breast cancer are caused by osteoclast stimulation. It is mobilised by the increased levels of receptor activator of nuclear factor–κB ligand and inflammatory cytokines.^30-32^ High activities of osteoclasts are observed in the osteoporotic bone where tissue resorption is accelerated.^21,33^ Thus, the mechanism of osteoporosis fundamentally provides fertile premetastatic niches for tumour cells in bone.^34,35^ A clinical investigation showed the association of untreated osteoporosis with accelerated progression of bone metastasis, should it occur.^21^ Bone density is also an important risk factor in some cancers.^36-39^

The outcomes of adjuvant BMA use in postmenopausal women with eBC are controversial.^8-10,40^ These mixed results might be attributed to the classification of patients without considering bone density, although RCTs can logically adjust for unidentified confounding factors.^41-43^ Our results showed that the RO score was not an independent prognostic factor in postmenopausal women. It might due to the exclusion of the women diagnosed osteoporosis with adjuvant BMA use. Future studies should investigate the association between osteoporosis and BM in postmenopausal women with eBC. The present study also implies a substantial benefit of adjuvant use in patients with low bone density diagnosed with solid or liquid cancers causing metachronous bone lesions.

From the perspective of retrospective studies with use of machine learning, our study design provides a new one, uncovering new risk factors and hidden at-risk populations. We regarded the output from the deep learning application as exposure, whereas most previous studies evaluated output from deep learning. We considered output as a patient characteristic, uncovering an association between the output and clinically crucial endpoints. We utilized the DL application, OPSCAN, osteoporotic precise screening using chest radiography and an artificial neural network. It was developed using 48353 chest radiography correspondence to T-scores.^17^ As the patient age for the matched data was 20 years or older, those with osteoporosis, osteopenia, and a normal range of T-scores were widely included. Furthermore, the model’s efficacy was confirmed by RCTs which aimed to identify individuals at high risk of osteoporosis.^16^

Osteoporosis must be viewed as a pathological condition which affects the entire skeleton,^44^ irrespective of whether it is defined by DXA of the lumbar spine or assessment of RO is performed by chest radiography. Such examinations of a specific skeletal location can limit the ability to determine the state of the entire skeleton. Further studies using whole-body computed tomography can reveal the heterogeneity of skeletal status and new aspects of its association with BM.

This study has some limitations that warrant further consideration. First, it was a retrospective study. Selection and recall biases may have affected our results. Second, the risk of osteoporosis due to machine learning was not strictly consistent with the definition of the WHO criteria,^45^ although the adopted model was trained and developed using over 48000 images in which the patient age for the matched data was 20 years or older.

## Conclusion

The risk of osteoporosis was identified as an independent prognostic factor of bone metastasis in women with eBC. A clear association between the risk of osteoporosis and bone metastasis in premenopausal women with eBC was observed. Using deep learning technology, we identified a poorly cared-for population from the perspective of the bone environment: premenopausal women with eBC, for whom we suggest an expanded indication for the adjuvant use of BMAs.

## Data Availability

All data produced in the present study are available upon reasonable request to the corresponding authors

## DECLARATIONS

### Consent for publication

Not applicable.

### Availability of data and materials

Data from this study, including those from individual participants, are not available for sharing. Summary statistical data are available from the corresponding author upon reasonable request.

### Authors’ contributions

All the authors contributed to the manuscript and approved the submitted version. After unmasking the outcome data, all authors had the final responsibility for the decision to submit it for publication. KT, AI, KW, and MT verified the raw data used in this study. HS conceptualised this study. HS and NT contributed to the methodology and formal data analysis. HS was responsible for the data analysis software used. All the authors validated the findings of the study. HS contributed to the formal data analysis. CK and TO conducted experiments. NT, KW, MY, and MT provided the necessary resources for this study. CK and TO curated data. HS wrote the original manuscript draft. NI and MT reviewed and edited the manuscript. HS created visualisations of the study findings. NI and MT supervised this study. TS was involved in project administration.

## Acknowledgements

Not applicable.

## References

1. Giaquinto AN, Sung H, Newman LA, et al: Breast cancer statistics 2024. CA Cancer J Clin 74:477–495, 2024

2. Heer E, Harper A, Escandor N, et al: Global burden and trends in premenopausal and postmenopausal breast cancer: a population-based study. Lancet Glob Health 8:e1027–e1037, 2020

3. Fabiano V, Mandó P, Rizzo M, et al: Breast Cancer in Young Women Presents With More Aggressive Pathologic Characteristics: Retrospective Analysis From an Argentine National Database. JCO Global Oncology:639–646, 2020

4. Pan Y, Lin Y, Mi C: Clinicopathological characteristics and prognostic risk factors of breast cancer patients with bone metastasis. Ann Transl Med 9:1340, 2021

5. Yao YB, Zheng XE, Luo XB, et al: Incidence, prognosis and nomograms of breast cancer with bone metastases at initial diagnosis: a large population-based study. Am J Transl Res 13:10248–10261, 2021

6. Patanaphan V, Salazar OM, Risco R: Breast cancer: metastatic patterns and their prognosis. South Med J 81:1109–12, 1988

7. Zhang W, Bado IL, Hu J, et al: The bone microenvironment invigorates metastatic seeds for further dissemination. Cell 184:2471-2486.e20, 2021

8. Gnant M, Pfeiler G, Steger GG, et al: Adjuvant denosumab in postmenopausal patients with hormone receptor-positive breast cancer (ABCSG-18): disease-free survival results from a randomised, double-blind, placebo-controlled, phase 3 trial. The Lancet Oncology 20:339–351, 2019

9. Gnant M, Frantal S, Pfeiler G, et al: Long-Term Outcomes of Adjuvant Denosumab in Breast Cancer. NEJM Evid 1:EVIDoa2200162, 2022

10. Adjuvant bisphosphonate treatment in early breast cancer: meta-analyses of individual patient data from randomised trials. The Lancet 386:1353–1361, 2015

11. Rachner TD, Coleman R, Hadji P, et al: Bone health during endocrine therapy for cancer. Lancet Diabetes Endocrinol 6:901–910, 2018

12. Moretti L, Richelmi L, Cosentini D, et al: Adjuvant denosumab for early breast cancer–Evidence and controversy. The Breast 78:103826, 2024

13. Fan Y, Li Q, Liu Y, et al: Sex- and Age-Specific Prevalence of Osteopenia and Osteoporosis: Sampling Survey. JMIR Public Health Surveill 10:e48947, 2024

14. Shariati-Sarabi Z, Rezaie HE, Milani N, et al: Evaluation of Bone Mineral Density in Perimenopausal Period. Arch Bone Jt Surg 6:57–62, 2018

15. Chawla J, Sharma N, Arora D, et al: Bone densitometry status and its associated factors in peri and post menopausal females: A cross sectional study from a tertiary care centre in India. Taiwanese Journal of Obstetrics and Gynecology 57:100–105, 2018

16. Lin C, Tsai D-J, Wang C-C, et al: Osteoporotic Precise Screening Using Chest Radiography and Artificial Neural Network: The OPSCAN Randomized Controlled Trial. Radiology 311:e231937, 2024

17. Tsai DJ, Lin C, Lin CS, et al: Artificial Intelligence-enabled Chest X-ray Classifies Osteoporosis and Identifies Mortality Risk. J Med Syst 48:12, 2024

18. Ryu J, Eom S, Kim HC, et al: Chest X-ray-based opportunistic screening of sarcopenia using deep learning. J Cachexia Sarcopenia Muscle 14:418–428, 2023

19. Young EM, Farmer JD: Preoperative Chest Radiography in Elective Surgery: Review and Update. S D Med 70:81–87, 2017

20. Eisen A, Somerfield MR, Accordino MK, et al: Use of Adjuvant Bisphosphonates and Other Bone-Modifying Agents in Breast Cancer: ASCO-OH (CCO) Guideline Update. Journal of Clinical Oncology 40:787–800, 2022

21. Chen H-M, Chen F-P, Yang K-C, et al: Association of Bone Metastasis With Early-Stage Breast Cancer in Women With and Without Precancer Osteoporosis According to Osteoporosis Therapy Status. JAMA Network Open 2:e190429–e190429, 2019

22. Guise TA: Bone loss and fracture risk associated with cancer therapy. Oncologist 11:1121–31, 2006

23. Saleh K, Carton M, Dieras V, et al: Impact of body mass index on overall survival in patients with metastatic breast cancer. Breast 55:16–24, 2021

24. Chiu C-T, Lee J-I, Lu C-C, et al: The association between body mass index and osteoporosis in a Taiwanese population: a cross-sectional and longitudinal study. Scientific Reports 14:8509, 2024

25. Fane ME, Chhabra Y, Alicea GM, et al: Stromal changes in the aged lung induce an emergence from melanoma dormancy. Nature 606:396–405, 2022

26. Turrell FK, Orha R, Guppy NJ, et al: Age-associated microenvironmental changes highlight the role of PDGF-C in ER+ breast cancer metastatic relapse. Nature Cancer 4:468–484, 2023

27. Møller AMJ, Delaissé J-M, Olesen JB, et al: Aging and menopause reprogram osteoclast precursors for aggressive bone resorption. Bone Research 8:27, 2020

28. Yang Q, Wei Z, Wei X, et al: The age-related characteristics in bone microarchitecture, osteoclast distribution pattern, functional and transcriptomic alterations of BMSCs in mice. Mechanisms of Ageing and Development 216:111877, 2023

29. Chung PL, Zhou S, Eslami B, et al: Elect of age on regulation of human osteoclast dilerentiation. J Cell Biochem 115:1412–9, 2014

30. Nguyen DX, Bos PD, Massagué J: Metastasis: from dissemination to organ-specific colonization. Nature Reviews Cancer 9:274–284, 2009

31. Mundy GR: Metastasis to bone: causes, consequences and therapeutic opportunities. Nat Rev Cancer 2:584–93, 2002

32. Roodman GD: Mechanisms of bone metastasis. N Engl J Med 350:1655–64, 2004

33. Takegahara N, Kim H, Choi Y: Unraveling the intricacies of osteoclast dilerentiation and maturation: insight into novel therapeutic strategies for bone-destructive diseases. Experimental & Molecular Medicine 56:264–272, 2024

34. Talmadge JE, Fidler IJ: AACR centennial series: the biology of cancer metastasis: historical perspective. Cancer Res 70:5649–69, 2010

35. Weilbaecher KN, Guise TA, McCauley LK: Cancer to bone: a fatal attraction. Nat Rev Cancer 11:411–25, 2011

36. Trabert B, Geczik AM, Bauer DC, et al: Association of Endogenous Pregnenolone, Progesterone, and Related Metabolites with Risk of Endometrial and Ovarian Cancers in Postmenopausal Women: The B∼FIT Cohort. Cancer Epidemiol Biomarkers Prev 30:2030–2037, 2021

37. Dallal CM, Lacey JV, Jr., Pfeiffer RM, et al: Estrogen Metabolism and Risk of Postmenopausal Endometrial and Ovarian Cancer: the B ∼ FIT Cohort. Horm Cancer 7:49–64, 2016

38. Dallal CM, Tice JA, Buist DS, et al: Estrogen metabolism and breast cancer risk among postmenopausal women: a case-cohort study within B∼FIT. Carcinogenesis 35:346–55, 2014

39. Dallal CM, Brinton LA, Bauer DC, et al: Obesity-related hormones and endometrial cancer among postmenopausal women: a nested case-control study within the B∼FIT cohort. Endocr Relat Cancer 20:151–60, 2013

40. Coleman R, Finkelstein DM, Barrios C, et al: Adjuvant denosumab in early breast cancer (D-CARE): an international, multicentre, randomised, controlled, phase 3 trial. The Lancet Oncology 21:60–72, 2020

41. Pourhoseingholi MA, Baghestani AR, Vahedi M: How to control confounding elects by statistical analysis. Gastroenterol Hepatol Bed Bench 5:79–83, 2012

42. Vetter TR, Mascha EJ: Bias, Confounding, and Interaction: Lions and Tigers, and Bears, Oh My! Anesth Analg 125:1042–1048, 2017

43. Nair B: Clinical Trial Designs. Indian Dermatol Online J 10:193–201, 2019

44. Pietschmann P, Peterlik M: [Pathophysiology of osteoporosis]. Wien Med Wochenschr 149:454–62, 1999

45. LeBoff MS, Greenspan SL, Insogna KL, et al: The clinician’s guide to prevention and treatment of osteoporosis. Osteoporos Int 33:2049–2102, 2022

